# Exploring Perceptions and Experiences of ChatGPT in Medical Education: A Qualitative Study Among Medical College Faculty and Students in Saudi Arabia

**DOI:** 10.1101/2023.07.13.23292624

**Authors:** Noura Abouammoh, Khalid Alhasan, Rupesh Raina, Khalid A. Malki, Fadi Aljamaan, Ibraheem Tamimi, Ruaim Muaygil, Hayfaa Wahabi, Amr Jamal, Jaffar A. Al-Tawfiq, Ayman Al-Eyadhy, Mona Soliman, Mohamad-Hani Temsah

**Author notes:** Corresponding authors: Rupesh Raina and Mohamad-Hani Temsah. Author Contributions: Noura Abouammoh and Mohamad-Hani Temsah roles were conceptualisation, data curation, formal analysis, funding acquisition, investigation, methodology, project administration, resources, software, supervision, validation, visualization, writing – original draft, and writing – review & editing the final version. Both authors directly accessed and verified the underlying data reported in the manuscript. Khalid Alhasan, Khalid H Malki, Ibraheem Altamimi, Ruaim Muaygil, Hayfaa Wahabi, Amr Jamal, Mona Soliman, Jaffar A. Al-Tawfiq, and Ayman Al-Eyadhy contributed to the data curation, investigation, methodology, resources, software, validation, visualization, writing – original draft, and writing – review & editing the final version. All authors have read and agreed to the published version of the manuscript. Funding: This research received no external funding. **Data Availability Statement:** The deidentified participant data collected for this study will be made available to others, upon reasonable request from the corresponding authors, with investigator support, after approval of a proposal, in agreement with the IRB-provided signed data sharing agreement.

## Abstract

**Background:** With the rapid development of artificial intelligence (AI) technologies, there is a growing interest in the potential use of AI-based tools like ChatGPT in medical education. However, there is limited research on the perceptions and experiences of faculty and students with ChatGPT, particularly in Saudi Arabia.

**Objective:** This study aimed to explore the knowledge, perceived benefits, concerns, and limitations of using ChatGPT in medical education, among faculty and students at a leading Saudi Arabian university.

**Methods:** A qualitative study was conducted, involving focused meetings with medical faculty and students with varying levels of ChatGPT experience. A thematic analysis was used to identify key themes and subthemes emerging from the discussions.

**Results:** Participants demonstrated good knowledge of ChatGPT and its functions. The main themes were: (1) knowledge and perception of ChatGPT, and (2) roles of ChatGPT in research and medical education. The perceived benefits included collecting and summarizing information and saving time and effort. However, concerns and limitations centered around the potential lack of critical thinking in the information provided, the ambiguity of references, limitations of access, trust in the output of ChatGPT, and ethical concerns.

**Conclusions:** This study provides valuable insights into the perceptions and experiences of medical faculty and students regarding the use of ChatGPT in medical education. While the benefits of ChatGPT were recognized, participants also expressed concerns and limitations requiring further studies for effective integration into medical education, exploring the impact of ChatGPT on learning outcomes, student and faculty satisfaction, and the development of critical thinking skills.

## Introduction

Artificial intelligence (AI) is a computer-based technology invented as a digital system to imitate and expand human intellect and skills. The wide use of AI technology has changed the medical field considerably towards more efficient patient’s management. As an example, integration of AI in diagnostic modalities in addition to introduction of machine based surgical treatment such as robotic surgery, have effectively promoted diagnostic accuracy and treatment with saved healthcare professional’s workload(1-3). All these changes that were integrated into the medical practice, in addition to hundreds if not thousands of future applications of AI in medical practice, need to be accompanied by changes in the medical teaching and training curricula. AI technology integration in medical education and medical research will not only contribute to patients’ care but potentially will improve if not revolutionize the medical education system(4, 5). AI’s rapid involvement in medical education generated significant interest among educators and researchers recently (6-9).

One of the pioneer and popular AI-based tools is ChatGPT, a language model developed by OpenAI that uses natural language processing to generate human-like responses to queries(10). ChatGPT has the potential to enhance medical education by providing an alternative and efficient means of accessing information. A recent study showed that 76.7% believed ChatGPT could positively impact the future of healthcare systems(11). However, little is known about the perceptions and experiences of faculty and students/trainee of ChatGPT in the context of medical education within Saudi Arabia.

The healthcare sector in Saudi Arabia is experiencing dramatic growth and reformatting, with a strong emphasis on prioritizing medical education. Therefore, such a healthcare system could benefit from a thorough strategy with substantial investments in medical, nursing, and other specialized educational disciplines(12). As medical education evolves, the use of AI-based tools like ChatGPT could potentially transform the way medical education is delivered in the region(13). Therefore, it is crucial to explore the medical faculty staff and students’ knowledge, perceived benefits, concerns, and limitations of ChatGPT application in medical education.

This qualitative study seeks to explore the knowledge base of new AI chatbots, like ChatGPT, utilization in medical education. Through gaining more understanding of the potential advantages and limitations of ChatGPT, this research can provide valuable insights to shape the creation of successful approaches for integrating AI-based tools into medical education in Saudi Arabia and similar medical education systems in the most efficient way.

## Methods

This study was conducted using a focus group technique at the College of Medicine, King Saud University, a leading university in Saudi Arabia(14). The study included faculty and students from different levels.

Participants were recruited from the College of Medicine through purposive sampling. The sample included six medical faculty members (two associate professors and four professors), and six medical students (two 2^nd^ year, two 3^rd^ year, one 4^th^ year and one 5^th^ year), with varying levels of experience with ChatGPT. Two focus group discussions were conducted in April 2023 on Zoom platform, one with faculty members and the other with students, and each group consisted of six participants. The discussions were conducted in the English language. Two of the authors served as moderators, each discussion lasted for approximately one hour. To ensure accuracy, the discussions were audio-recorded and transcribed word-for-word.

The assessed topics were as of the following: participants’ familiarity with the recently launched ChatGPT, its uses, facilitators, and hinderers of its incorporation in medical education. Probs and follow-up questions were allowed depending on participants’ responses. Themes were saturated after the second interview. Thematic analysis was used to analyze the data using priori themes and allowing new themes to emerge from the data. The transcripts were read multiple times to identify patterns and themes that emerged from the data. A coding framework was developed based on the research questions and applied to the data using NVivo 12 software(15). Themes were identified and refined through an iterative process of coding, reviewing, and discussing the data among the research team until consensus was reached.

Ethical approval was obtained from the Institutional Review Board at the participating university (Ref. No. 23/0155/IRB). Verbal informed consent was obtained from all participants prior to their inclusion in the study and for the recording. Participants were informed about the study’s purpose, the voluntary nature of their participation, and their right to withdraw at any time. Pseudonyms were used to protect the anonymity of the participants.

## Results

Six medical faculty staff and six medical students with different experience with ChatGPT participated in the study. Table 1 shows their demographic data.

**Table 1:** Participants position, department, and frequency of ChatGPT use:

| Participant Code | Position | Department | Using Chat GPT |
| --- | --- | --- | --- |
| P1 | Faculty | Critical care department,<br>College of medicine | Regular user |
| P2 | Faculty | ENT department,<br>College of medicine | Regular user |
| P3 | Faculty | Family medicine,<br>College of medicine | Not a user yet but heard<br>and read about<br>ChatGPT |
| P4 | Faculty | Pediatrics department,<br>College of medicine | Not a user yet but heard<br>and read about<br>ChatGPT |
| P5 | Faculty | Pediatrics department,<br>College of medicine | Regular user |
| P6 | Faculty | Medical Education<br>Department,<br>College of medicine | Regular user |
| S1 | Student | College of medicine | Regular user |
| S2 | Student | College of medicine | Regular user |
| S3 | Student | College of medicine | Regular user |
| S4 | Student | College of medicine | Regular user |
| S5 | Student | College of medicine | Regular user |
| S6 | Student | College of medicine | Regular user |

Analysis of the data from discussion generated two main themes: 1. Participants’ perception of ChatGPT. 2.ChatGPT utilization in medical education and research. Figure 1 display the thematic framework of Participants’ perception about the use of ChatGPT in general and in medical education.

**Figure 1:**
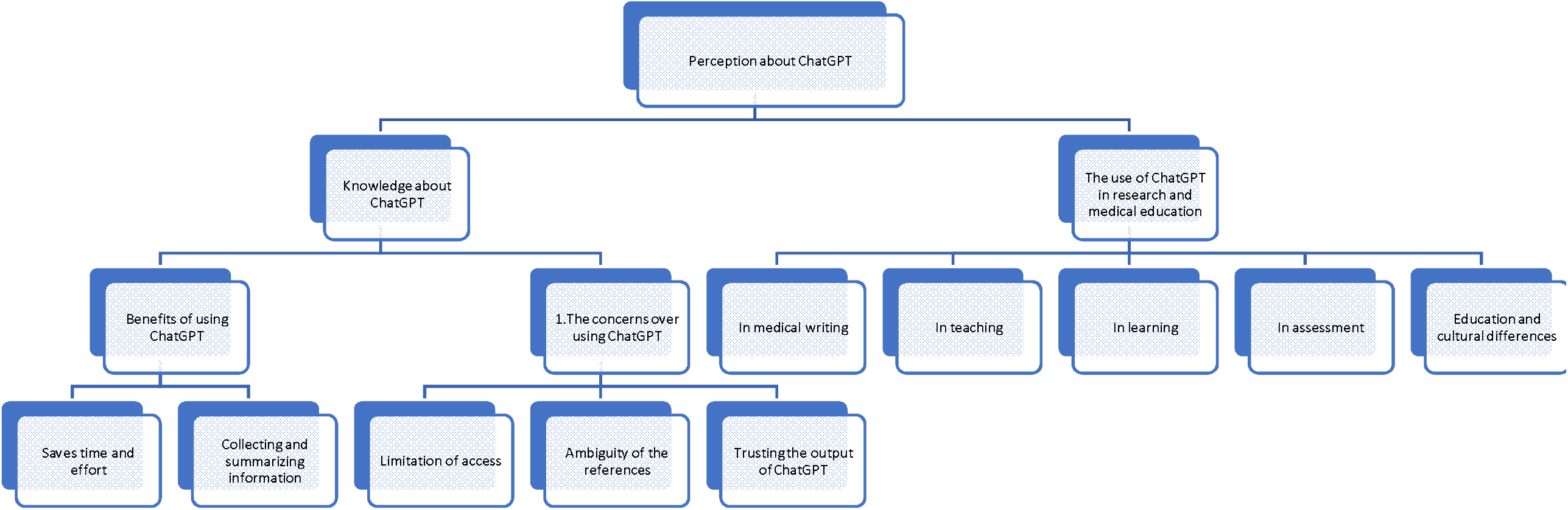
Thematic framework of participants’ perception on using ChatGPT.

### 1. Participants’ perception of ChatGPT

All participants showed good knowledge about ChatGPT nature, and they agreed on the main function and goal from ChatGPT. For example, one participant noted:

> *“The idea from this software is that it will chat with you regarding any topic you will ask about…it chats with me in a human like manner, and collect for me the answers from all over these resources, and display them for me” P1*

One student described Chat GPT as an “assistant” and another one explained:

> *“Artificial intelligence helps you execute the command that you asked to be executed” S2 And*
>
> *“It’s another way of searching for information that’s highly accurate*… *depending on what you search for and how you search for it” S5*

The resources of information in ChatGPT were discussed among the group compared with other traditional search engines. While most participants were not disturbed by the sources, two faculty participants had different views.

> *“Other databases like Google, are doing the same. It’s the same data retriever” P3* Compared to,
>
> *“It is not at all Google, at all, and to prove that; Google starts trying to also generate a chat platform similar to ChatGPT” P2*

The following present participants’ views on perceived benefits and concerns of using ChatGPT.

### 1.1 Benefits of using ChatGPT

Participants used words such as “beautiful” and “comprehensive” to describe aspects of using ChatGPT. Two main subthemes emerged from the focus group discussions as benefits of using ChatGPT.

#### 2.2.3 Collecting and summarizing information

Most faculty participants believed that searching for information through ChatGPT is more beneficial than using the standard search engines, as the former saves time by summarizing and textualizing the raw information output from the search.

> *“It (ChatGPT) is beautiful in collecting information and presenting it as a simplified text that can be easily digested and understood” P2*

Another faculty added,

> *“It will search it (information) for you; it will critically appraise it and give you the final result” P1* Students shared the same view:
>
> *“I find it (ChatGPT) to be more directive towards what you ask, and to the point, mostly because when you look for something on a normal search engine, such as Google…you have to go into some sub web pages which has an answer and look between all the thousands of answers to find one. While ChatGPT will give it to you concisely like this is option A, option B, option, C” S3*

However, despite the former positive point mentioned, one faculty participant was more conservative in her comments about using AI in collecting data. Unlike the view of the previous participant, another participant does not perceive the information displayed by ChatGPT as critically appraised. The participant noted:

*“The problem of collecting all the information in one place is that collecting the information and giving it in a nutshell, in one place. This machine is not critically thinking” P3*

#### 1.1.2 Saves time and effort

While most participant agreed on the kind of information given by ChatGPT and on how to obtain this information, opinions varied in terms of whether using ChatGPT saves time and efforts.

One faculty mentioned:

> *“It saves time when I’m stock in generating exam question” P2 A student added:*
>
> *“it’s not accurate, but at least it saves your time. This is the most important point” S4*

On the other hand, another faculty participant subtly disclosed her denunciation on the functionality of ChatGPT. She believed that ChatGPT helps partly in performing tasks, but that advantage does not outweigh the caution of dealing with the information generated from it, she is going to put some effort anyway.

> *“Me as a researcher. When I search for information I’m putting it together, it tries to put it for me. So far, I can’t see it as superior to the human mind” she added “it does some of the work for you, but you have to take it with a bunch of salt” P3*

### 1.2 Concerns over using ChatGPT

When the participants discussed the drawbacks of using ChatGPT, they mentioned words such as “hallucination”, “blinding euphorically” and “caution” to describe their experiences in using ChatGPT. The following subthemes emerged as perceived drawbacks of ChatGPT.

#### 2.2.3 Referencing unreliability

Although most participants thought that the internet is the source of ChatGPT information:

> *“It’s the same data retriever as Google” P3*

Other participants had an opposing view.

> *“Article references, and the citation for those references has to be taken with caution” P2*

Faculty experienced situations where they doubted the reference of the information provided by ChatGPT. For example, one faculty noted:

> *“I’m not sure what are the sources used to develop the information… It will just mention differences in views, even if you ask for references, it will not mention them…or at least it will not volunteer to mention them” P1*

Another participant defended that and replied:

> *“If it doesn’t have access to the reference, it will tell you I don’t have access, but if the book is online, it can refer to that” P2*

Similarly, a student commented:

> *“It has multiple resources rather than searching for a website that have the answer. It gives you the answer and references to these websites” S5*

One participant suggested that the referencing unreliability is evidence supporting her view of not relying on ChatGPT, she noted:

> *“The problem with the references is just a manifestation of what is under this” P3*

#### 1.2.2. ChatGPT access to information

Some participants acknowledged the limitation of ChatGPT in accessing all information available online. One participant explained:

“One of the restrictions regarding medical search that it’s restricted to like PubMed… there are some other medical websites that it cannot access yet” P4

Because of that, participants encouraged using ChatGPT with caution. One faculty noted: *“We don’t know the algorithm behind the search nor exactly how it looks for information” P5*

Student participants explained that ChatGPT is designed by human, thus its restricted to what it was coded to do and search. One noted:

> *“it’s not free of bias. If you’re asking it to answer something morally wrong or something illegal. It will not answer you because it is constrained. So, it is not fully free from human constraints” S3*

#### 1.2.3 Trusting the output of ChatGPT

All participant believed that users of ChatGPT should not fully trust the information it displays, and some encouraged using it with “caution” while others encouraged using it for new topics as a jumpstart.

> *“You should not take it (information from ChatGPT) for granted; you have to review what’s there, but it gives you a nice idea, very excellent ideas…* It shed lights on some certain angles that you are not looking for*” P2*

A student added:

> *“It’s a tool! …it is continuously improving and getting updated” S5*

According to the participants, trusting information from ChatGPT depends on your previous knowledge about the topic searched. One participant explained:

“You have to have the ability to differentiate between what is reliable and what is not reliable… Myself, I am not well versed in medical education. For example, I am well invested in research, I would take whatever it is giving me on medical education, but I can filter information regarding research and judge it well” P3.

One student agreed with the former point:

> *“Depends on what you’re asking. Sometimes it’s very accurate. Sometimes it’s not…But as a human being you have an idea about what you’re asking for so you can say if its accurate or if you should doubt the answer” S4*

The unfamiliarity of ChatGPT users about the methodology of its search added to the trust issue. A faculty explained:

> *“If I know whether this methodology (ChatGPT searching methodology) is a scientific methodology, how to search for the papers, how to extract the information from the paper, how it appraises it. What are the sources that this engine has access to. That will augment the reliability of the experience.” P1*

One participant mentioned that ChatGPT; cannot be used for critical thinking in certain contexts thus it cannot be fully trusted:

> *“It cannot give you what is relevant to your community? What is relevant to your population. What is relevant to your students… We are prioritizing this thing (ChatGPT) over human intellect!” P3*

Another faculty participant was defendable in arguing the above presented view by noting:

> *“If you ask ChatGPT about something in geology for example, it will start with ‘I am not a geologist’ and then move on with the dialogue… and it finishes the response by ‘it is very important to refer to those sources’” P2*

Faculty participants raised an ethical concern that may affect trusting ChatGPT as a source of information. One participant explained:

> *“Can drug company pay ChatGPT to display answers that are in favour of certain medication? Could ChatGPT be manipulated? They are sure looking for money somehow!” P1*

## 2. ChatGPT utilization in medical education and research

Participants discussed the use of ChatGPT in medical education from three aspects: learning, teaching and assessment.

### 2.1 In learning

Most faculty support the importance of utilizing ChatGPT in the process of learning because:

> *“Students are no longer enjoying the usual long lectures, or didactic lectures but they enjoy more challenging aspects exploring a new experience, and living it*… *I think the ChatGPT could be used as a very good trigger for the students to go and read and find out more, discuss among themselves and go explore this with their seniors, with their educators” P5*

One faculty participant confirmed the prospect of using ChatGPT to recall facts but not for opinions and debates. She noted:

> *“If you use it (ChatGPT) just for recalling then no problem… But if you want to make inferences you should not use it” P3*
>
> *Most student participants did not use it to obtain information. One student said:*

“I’m not seeing it as a search engine. I don’t look up medical information on it, or anything, because I find the classic search engines easier” S2

Because “I know exactly where the reliable sources are. Then I am able to take the information with confidence” S1

Student also added,

> *“If I am going to write a sentence like, what is the regular or the normal range of glucose? as I start to write this question on Google, it will be on the suggestion before I finish my writing. Not like ChatGPT. I have to write the whole question” S1*

A faculty participant raised the concern of students and faculty losing their critical thinking skills if they depended on ChatGPT, she explained:

“It is dangerous…because we are replacing critical thinking. We are prioritizing this thing over human intellect” P3

A student added,

> *“If I’m someone who never wrote any literature, and I’m only using chatGPT exclusively. I can see it impacting my writing skills heavily” S2*

One faculty, however, did not support using ChatGPT to list facts because:

> *“Because we don’t know the source. We don’t know the methodology of getting this information” P5*

Some participants thought that ChatGPT cannot be used in medical related information and decision making as it does not give the source of evidence. One of the faculty participants, despite supporting the use of ChatGPT in general, noted:

> *“If you look at the other search engines for which support medical information, they present like up-to-date information…ChatGPT is very complex, and the methodology and the algorithm it uses is not clear so, it is not a reliable source of information for decision making and for serious information” P1*

Other participants debated that the accuracy of the information displayed depend on the searching skills of person looking for it:

> *“Prompt questions will make the difference in getting the response, and I recommend digging into the prompts to get more accurate answers and doing this is important to acquire the right answer” P2*

“You get the response according to the precision of the search” S1 The issue of updated sources in ChatGPT was also raised:

> *“We need to be cautious about using the information… the medical field information is changing very quickly, so we have to be careful about this point” P4*
>
> Another faculty added that it should be used to get an idea about a topic, but further reading is important for students:
>
> *“ChatGPT is like a short fast access to a topic, it helps to get the most important information… they (students) need to read the references” P1*

In general, students do not resort to ChatGPT as the first source for new information. However, they use it to confirm their knowledge about a topic as a “collateral resource”.

### 2.2 In teaching and training

Some participants believed that teaching modalities should change after the introduction of AI. One faculty noted:

> *“Students don’t need that large, enormous amount of information. They can get this now its easily accessed to anyone at any time” P4*

All faculty participants realized that the presence of AI should shift the teaching manner from memorization to critical thinking as the later cannot be provided by AI:

> *“We have to invest more in the skills of our medical students and problem-solving critical thinking analysis. These are the areas that is lacking in the ChatGPT, and that we need to focus more on” P4*

One participant refrained from sending students to search for information on ChatGPT because:

> *“Do we have the awareness to differentiate between what the students have learned and what the ChatGPT has given them?” P3*

Furthermore, students may not attend the lecture if they could find an alternative source of information:

> *“I’m not coming to the lecture. Why are you not coming to the lecture? All the information you are giving me, and 90% more I can get from ChatGPT” P3*
>
> Another concern raised from one faculty regarding the lecturers and trainers: *“Have our faculty have enough knowledge to use and recommend ChatGPT?” P3 However, students did not see themselves relying on ChatGPT for learning:*
>
> *“We just need to be familiar on how to use ChatGPT and use it as a tool that support our search rather than completely relying on it” S2*

Faculty participants differentiated between postgraduate and undergraduate students’ need and use of ChatGPT. One faculty (P3) mentioned that using AI in training postgraduates would be difficult because it depends on building skills while in undergrad it depends on memorization. Another faculty argued: *“Maybe this is the current situation. But t’s time to change this. It shouldn’t be memorization” P4*

Using ChatGPT could save medical educators’ time and effort to create problems to be utilized for discussion with the students,

> *“One problem would take weeks from our team and long hours of sitting together and creating the problems that we teach in the problem-based learning sessions. So, it would be really interesting to see how ChatGPT deals with this” P4*.

One of the participants expressed her worries about utilizing ChatGPT in teaching and training. Her justification was the lack of training on:

> *“Humanity and the communication, the teamwork. All of this” P4* Another faculty added:
>
> *“It (ChatGPT) will never be a teacher or a trainer, and the medicine in general is not just memorizing” P1*

### 2.3 In assessment

Three faculty participants mentioned using ChatGPT in the assessment. One faculty mentioned that he used ChatGPT to generate exam questions:

> *“I asked ChatGPT to generate questions for me with scenario and without scenario… it was good to Very good. It’s not reaching to excellent level. I have to review and modify.” P2*
>
> Another faculty suggested using ChatGPT to create problems to be solved at bedside teaching.
>
> *“Create like quizzes, quick, short answer questions that will make the lectures or the tutorials more interactive” P4*

Faculty participant (4) suggested Using ChatGPT to review the assessment questions. If ChatGPT answered the assessment questions correctly and gained a high score, that means the assessment contents were based on the objective of knowledge and memorization and not on critical thinking. Some participants used ChatGPT for to review papers or provide new idea about a paper.

> *“I used it to study limitations of studies and the future recommendations for studies I was asked to review … it gives me ideas” P2*

Cheating and plagiarism was one of the concerns raised by the faculty during the discussion:

> *“We have to be very careful about the cheating… the misuse of the ChatGPT from our medical students in medical assignments” P4*

In line with the former comment, one student explained how he uses ChatGPT in writing assignments:

> *“I mainly use it to write something, and then I just review it and edit it…mainly in research or some essays…I’d give it some data, and I ask it to write a paragraph that summarizes this data, for example, or an introduction to something for example (Disease X). And it will do so. I see variable results sometimes that I think is very, very good, sometimes it’s all over the place, but for the most part it’s very good” S2 The same student added:*
>
> *“If I use ChatGPT to write a homework assignment. Am I cheating or not? I think that’s an issue that has to be resolved” S2*

Another student used it to help in staying within the word limit:

> *“When I write an essay or something, and I go over the word limit. I sometimes try to use ChatGPT with a decent success, to shorten it” S3*
>
> *Others use it to collect resources:*
>
> *“It can make your job way easier. For example, if you have a research assignment to just collect the resources on this topic” S3*

Despite all of the above-mentioned arguments about the use of ChatGPT in medical education, all participants decided to be open minded to accept it. For example:

> *“I think it’s coming in the near future, and we need to live in the reality to adjust and take the best out of it” P4*

## Discussion

The participants demonstrated a good understanding of ChatGPT and its functionalities, some described it as an “assistant” or a highly accurate information search tool. They highlighted differences in their opinions regarding ChatGPT’s resources compared to traditional search engines like Google. In one study comparing both platforms, while experts generally considered ChatGPT-generated responses reliable and useful, some considered them dangerous, with 40% concluding that ChatGPT responses were perceived as more valuable than Google (16). Google operates by scanning billions of web pages, indexing the content, and ranking it according to relevance before presenting users with a list of links to browse (17). In contrast, ChatGPT provides a more user-appealing and faster solution for busy users, by delivering a single, synthesized answer based on its own search and algorithms, acquiring its conversational abilities and knowledge by being trained on millions of websites, as long as the information was published before late 2021 (18). While few participants in our study found similarities between the two platforms, others argued that ChatGPT was distinct and that its launch even prompted several search engines to accelerate their developments of similar chat-based platforms. This is in line with other reported experts’ opinion about acknowledging the sophistication and nuances in the responses, but also recognizing that responses were frequently incomplete and sometimes misleading (16).

The perceived benefits of using ChatGPT in medical education were mainly centered around two subthemes: collecting and summarizing information and saving time and effort. Participants appreciated the ability of ChatGPT to provide a concise summary of search results, which they found more efficient than using standard search engines. They also noted the time-saving aspect of ChatGPT, particularly in generating exam questions or quickly providing information.

Despite the perceived benefits, participants raised concerns about potential limitations of ChatGPT in medical education. The lack of critical thinking, generation, lack of accuracy and critical appraisal of the information provided by ChatGPT was highlighted, emphasizing the importance of human judgment and critical thinking in this field(9). Participants suggested that while ChatGPT might be helpful in certain aspects of medical education, users should approach the information with caution and apply their own judgment. One study listed the following as possible disadvantages: lack of originality, inaccurate content, or unknown data sources(10). It is also uncertain how ChatGPT handles offensive material, false information, or plagiarism(19).

One study listed the following advantages of ChatGPT in medical education: automated scoring, teaching, and research assistance, and personalized learning(20). Another study listed enhanced personalized learning, critical thinking, and problem-based learning as advantages of ChatGPT in medical education(10). Banerjee et al reported that postgraduate trainee doctors have an overall positive perception of the impact of AI on clinical training(r). However, they found that AI will eventually reduce the trainees’ skills in ‘clinical judgement’ and ‘practical skills’(7).

The potential implications of using ChatGPT in medical education include improved efficiency, streamlined information gathering, and time-saving benefits. However, future research is needed to explore the impact of AI-based tools on medical learning and practice, such as the quality of education, student and faculty satisfaction, and the development of critical thinking skills. Ongoing research and evaluation are essential to ensure the effective integration of AI-based tools like ChatGPT into medical education while addressing potential concerns and limitations.

In addition, participants discussed concerns related to the ChatGPT’s referencing reliability, limitations in accessing specific medical websites or databases, trust of the information provided. Addressing these issues requires improvements in ChatGPT’s transparency, disclosure of citations algorithm allowing users to easily verify the information’s source and credibility, expanding access to resources. A previous study had cautioned that authors should be cautious with references when using ChatGPT(21).The limitations of ChatGPT in accessing specific medical websites or databases were also discussed, impacting its usefulness in medical education. To overcome these limitations, developers could work on expanding ChatGPT’s access to more resources and improving its search capabilities, ensuring a more comprehensive and reliable source of information.

Factors influencing users’ trust included their prior knowledge on the topic and unfamiliarity with ChatGPT’s search methodology. Emphasizing the importance of critical thinking and judgment when using ChatGPT as a source of information in medical education is crucial to ensure users are discerning and do not blindly trust the AI-generated content. For example, specific advice from ChatGPT were found at times to be incorrect in relation to cosmetic surgeries. That study found ChatGPT was 50% accurate for general cosmetic surgery and 80% accurate for rhinoplasty and blepharoplasty(22). However, a recent expletory review showed that ChatGPT has potential impact on medical education, scientific research, and medical writing(10). Another study showed concerns about ChatGPT advice in relation to antimicrobial stewardship. The appropriateness of duration varied; general course lengths were accurate, but source control was either incorrectly cited as justification for prolonging therapy or ignored entirely(23).

Ethical concerns, such as potential manipulation by pharmaceutical companies, were raised by participants. Maintaining transparency and integrity in AI-generated information is vital to address these concerns. Implementing measures such as third-party audits, strict guidelines, and continuous monitoring of ChatGPT’s information sources can help ensure the ethical use of ChatGPT in medical education. Ethical concern in addition to other concerns such as inability to reason beyond existing knowledge, and bias had also been reported previously(24).

Participants explored the potential of ChatGPT in generating exam questions and scenarios, enhancing bedside teaching, reviewing assessment questions, and addressing concerns of cheating and plagiarism. The discussion emphasized the need for faculty involvement in reviewing and modifying AI-generated content, as well as the importance of developing policies and strategies to tackle potential academic misconduct related to ChatGPT use. This particular issue should be considered as previous studies showed that ChatGPT had passed the AHA exam with 84% accuracy but failed the Taiwan’s family medicine exam and poorly on the urology self-assessment examination(25-27). Thus, the generated questions need to be carefully examined.

ChatGPT’s potential use in student assignments was also discussed, focusing on assisting in writing and editing, data analysis and gathering, and resource collection. Participants highlighted the need for students to critically review and modify the AI-generated content, ensuring that it aligns with academic standards and expectations. A recent review raised concerns about using ChatGPT for exam and assignment cheating by students(24). The cons of ChatGPT use in medical education had included loss of creativity and possible undermining of students’ capacity of critical thinking.

The discussions highlighted the importance of understanding the capabilities and limitations of each AI-driven tool, like ChatGPT, as well as the need for ongoing evaluation of their integration into medical education settings. One study showed that ChatGPT can answer first and second order questions in certain subjects such as microbiology(28).

Participants emphasized the importance of being open-minded and adapting new technologies like AI chatbots including ChatGPT. Addressing cultural differences in learning styles, ensuring relevance and accuracy of information, and encouraging local development and customization when incorporating ChatGPT into medical education across diverse cultural contexts and regions to better serve the needs of local students and educators.

In preparation for the future of medical education, educational institutions should be proactive in integrating AI technologies like ChatGPT into their curricula and teaching methodologies. This process should involve regular evaluations, ongoing improvements, and a strong emphasis on maintaining the essential human aspects of medical education, such as critical thinking, communication, and empathy.

## Strengths

One of the strengths of this study is the qualitative approach, which allowed for an in-depth exploration of participants’ experiences, perceptions, and concerns related to the use of ChatGPT in medical education. The focused meetings provided a platform for rich discussions among medical faculty and students, revealing diverse viewpoints and generating valuable insights into the potential benefits and challenges of integrating ChatGPT into medical education. Moreover, the study involved participants with varying levels of experience with ChatGPT, ensuring a comprehensive understanding of the perspectives of both novices and experienced users. The identification of themes and subthemes based on these discussions has laid a solid foundation for further research and exploration of AI-based tools like ChatGPT in the medical education context.

## Limitations

However, there are some limitations to the study. The sample size was relatively small, and the participants were primarily drawn from a single institution, which may limit the generalizability of the findings to other medical education settings. The study did not quantitatively assess the impact of ChatGPT on learning outcomes, satisfaction, or other measurable aspects of medical education, which could provide valuable data to supplement the qualitative findings. Additionally, since the study’s focus was on the opinions and experiences of faculty and students, the perspectives of other stakeholders, such as administrators and policymakers, were not captured. This limitation could affect the comprehensiveness of the study’s insights and the development of well-rounded strategies for the integration of ChatGPT into medical education. Furthermore, the study did not explore the long-term implications and potential changes in perception and usage of ChatGPT over time, as participants’ experience with the tool may evolve, altering their views on its benefits and limitations.

Future research should address these limitations by incorporating larger and more diverse samples from multiple institutions, as well as conducting quantitative studies to measure the impact of ChatGPT on various aspects of medical education. Additionally, incorporating the perspectives of other stakeholders, such as administrators and policymakers, could provide a more comprehensive understanding of the implications of ChatGPT in medical education. Longitudinal studies could be conducted to assess the changes in perception and usage of ChatGPT over time and evaluate the long-term effects of its integration into medical education.

## Conclusion

The findings of this study provide insights into the early perceptions and experiences of faculty and students with ChatGPT in the context of medical education in Saudi Arabia. Participants generally had good knowledge of ChatGPT, and perceived benefits such as timesaving and collecting and summarizing information. However, concerns were raised regarding the accuracy and critical appraisal of the information provided by ChatGPT, and the need for users to approach the information with caution. Other limitations, such as ambiguity of references and limitations of access to specific medical databases, were also noted.

This study highlights the need for ongoing research and evaluation to ensure that AI-based tools like ChatGPT are effectively integrated into medical education while addressing potential concerns and limitations. Educators and students must also maintain a strong foundation in critical thinking and judgment, and approach the information provided by ChatGPT with caution. As medical education continues to evolve, the integration of AI technologies like ChatGPT has the potential to transform the way medical education is delivered but must be done with a thoughtful and ethical approach.

## Data Availability

All data produced in the present study are available upon reasonable request to the corresponding author

## Acknowledgments

We have used ChatGPT, an AI-chatbot developed by OpenAI, to improve some readability and language of this work, without replacing researchers’ tasks. This was done with human oversight, and authors then carefully reviewed and edited the generated text, as we assure that the authors are ultimately responsible and accountable for the originality, accuracy, and integrity of their work. We would like to acknowledge the efforts in data curation in the focus groups, namely: Abdulaziz Alomar, Faisal Alomri, Hadi Alhemsi, Homoud Algadhib, and Ibrahim Alhezam. The authors extend their appreciation to the Deputyship for Research & Innovation, Ministry of Education in Saudi Arabia for funding this research (IFKSURC-1-3110).

